# Acceptability of receiving HIV and sexual and reproductive health services at hair salons among university students in Zambia: a cross-sectional survey

**DOI:** 10.64898/2026.08.06.26359879

**Authors:** Malena Chiaborelli, Lilian Nayame, Twaambo E. Hamoonga, Oliver Mweemba, Mamaswatsi Kopeka, Karen Hampanda, Alain Amstutz

## Abstract

Adolescent girls and young women in Zambia face barriers to accessing HIV and sexual and reproductive health services. We conducted a cross-sectional survey among 846 female students at a university in Zambia to assess acceptability of receiving these services in hair salons and explore whether HIV acquisition risk influenced acceptability of HIV services. Acceptability varied by service, ranging from 25% to 50%. Higher HIV acquisition risk may increase the acceptability for HIV services. Hair salons may be a promising community-based, demedicalized setting for delivering selected HIV and sexual and reproductive health services to HIV at-risk female students.

## Introduction

Adolescent girls and young women in eastern and southern Africa are one of the groups most affected by HIV due to biological, social, and structural factors. In 2023, they accounted for 63% of all new global HIV acquisitions, with more than 4,000 each week.^1^ In Zambia, HIV prevalence among women aged 20-24 years old is three times higher than among males of the same age group (6% vs 2%).^2^ Moreover, 29% of girls aged 15-19 have given birth, and unsafe abortions account for 15% of maternal deaths in the country.^3^

Several studies indicate challenges in the use of HIV prevention and sexual and reproductive health (SRH) services among students in higher learning institutions in Zambia.^4–6^ In a recent survey, 30% of sexually active female university students reported lack of any contraception use, and fewer than 1% used oral pre-exposure prophylaxis (PrEP), one of the most effective biomedical HIV prevention methods currently available in Zambia at no cost.^7^

Innovative approaches are needed to improve access, use, and continuation of essential SRH products, such as PrEP and modern contraceptive methods, among female university students in Zambia.^5,6^ Hair salons could serve as an innovative and easily accessible community setting to engage young women and help overcome certain barriers to SRH services. Groups from South Africa and Lesotho have explored the use of hair salons to deliver SRH education/services with promising findings.^8–10^ To our knowledge, no data from Zambia nor among African female university students is available on acceptability of hair salons as a venue for SRH education/services.

Therefore, we conducted a cross-sectional study to assess the acceptability of receiving HIV and other SRH services at hair salons among female students at a large public university in Lusaka, Zambia. We also aimed to explore whether HIV acquisition risk may influence the acceptability of receiving HIV and SRH services in this population.

## Methods

The study design was a cross-sectional survey derived from the recruitment questionnaire of a randomized trial assessing a mobile health peer navigator intervention on PrEP and contraception uptake and persistence among female university students in Zambia (NCT06852508).^11^ The study protocol was registered and is publicly available.^12^

The study was conducted at a large public university located in the capital city of Lusaka. The university provides on-campus HIV and SRH services and is surrounded by numerous nearby government-run and private health facilities and pharmacies. In Zambia, oral PrEP is available free of charge at all government health centers, while injectable PrEP is increasingly being offered in selected facilities.

Participants were eligible if they self-identified as women, were aged 18 to 25 years, and were enrolled as students at the university. We partnered with the university’s medical services and existing peer navigators on campus to distribute flyers with the survey QR code that led to a REDCap questionnaire. A virtual flyer was also distributed to student WhatsApp groups. After consent and screening questions, eligible participants were invited to complete the self-administered online survey. The questionnaire collected basic demographic information, nine behavioural items on objective epidemiologic risk of HIV acquisition using a validated HIV Risk Index, and eight questions assessing acceptability of receiving HIV/SRH services in hair salons (Supplementary 1 for the full questionnaire).^13^ All data were collected and stored in the REDCap database hosted on a secured server at the University of Colorado.

The sample size for this study was driven by the recruitment screening target for the overarching trial. No independent sample size calculation was conducted for this secondary analysis of the survey.

Survey data were analysed using descriptive statistics, including frequencies and percentages for categorical variables and medians with interquartile ranges (IQR) for continuous variables. Acceptability was assessed using five-point Likert-scale items asking participants whether they would feel comfortable receiving eight distinct HIV/SRH services from a hair stylist in a hair salon: HIV counselling, HIV self-testing, PrEP, family planning counselling, sexually transmitted infections (STI) counselling, condoms, birth control pills, and emergency contraceptive pills. Based on nine items, we calculated epidemiologic HIV risk.^11^ Each item contributed one point, resulting in a total score from 0 to 9. If any of the items was missing, we set the overall risk score to missing. Participants with three or more risk factors were classified as having high HIV acquisition risk, and all others as having low HIV acquisition risk.^11^ The threshold is based on prior research indicating that having three or more risk factors on the index is significantly associated with the WHO epidemiologic HIV high-risk threshold.^14^

We explored whether higher HIV acquisition risk may cause higher acceptability of receiving three specific HIV services at hair salons: HIV counselling, HIV self-testing, and PrEP. For each service, we fitted an ordinal logistic regression model, with the HIV risk index as the main exposure and acceptability of receiving the service as the outcome. Based on the hypothesized causal relationships (Directed Acyclic Graph available in the pre-specified and registered protocol), we considered age and financial situation as important potential confounders and adjusted all models for them.^12^ Data management and analysis were conducted using R version 4.5.1.

## Results

Between February and March 2025, 846 female students completed the survey. Baseline characteristics, HIV risk factors, and the proportion of participants with low and high HIV risk index are presented in Supplementary 2. Participants had a median age of 21 years (IRQ 20, 23). Economically, only 9.2% of participants reported their financial situation as comfortable, while the remaining students described their situation as less favorable. Regarding their living situation, most students lived either in a dormitory on campus (26.7%) or in a rented room around the campus (30.0%).

With respect to HIV acquisition risk, 49.9% of participants were currently in a sexual relationship, and 55.3% reported having at least one sexual partner in the past 12 months. Regarding knowledge of the HIV status of their last vaginal sex partner, 25.7% of participants said their partner had told them their HIV status, 17.3% reported that they had been tested together with their partner, while 11.3% had not asked their partner about it.

Overall, 19.5% of participants had sexual partners more than five years older, 6.6% had ever been pregnant, and 4.8% had had sex in exchange for money or gifts. Regarding perceptions of their partner’s sexual behavior, 6.6% of participants reported knowing that their partner had other sexual partners, 14.1% suspected it, and 17.8% were unsure. In the past six months, 13.2% of participants reported experiencing abnormal vaginal discharge or genital sores or ulcers, and 37.8% had had vaginal sex without a condom. According to the HIV risk index, 45.4% were classified as having high HIV acquisition risk with 3 or more reported risk factors.

In terms of acceptability of receiving HIV/SRH services in hair salons, reported levels varied between 25% to 50%, depending on the specific service (Figure 1 and Supplementary 3).

**Figure 1.** Acceptability of receiving HIV and SRH services in hair salons.

Acceptability was high for STI counselling (49%), HIV counselling (44%) and oral HIV self-testing (44%). In contrast, the highest lack of acceptability was reported for receiving birth control pill (58%), emergency contraceptive pill (53%), and PrEP (52%).

Having a higher HIV acquisition risk increased the odds of accepting oral HIV self-testing at the hair salon (adjusted OR: 1.15, 95% CI: 1.07 - 1.22) as well as PrEP (adjusted OR: 1.11, 95% CI: 1.04 - 1.18), while the result for HIV counselling was inconclusive (OR: 1.02, 95% CI: 0.95 - 1.09).

## Discussion

In this cross-sectional survey among over 800 female university students in Zambia, almost half were at increased risk of HIV acquisition. Acceptability of receiving HIV and SRH services in hair salons ranged from 25% to 50%, depending on the service. Our exploratory analysis suggested that higher HIV acquisition risk increases the acceptability of receiving HIV services in hair salons.

There are significant barriers to the accessibility of SRH services among female students in higher learning institutions in Zambia.^4^ For instance, there is favourable emergency contraception use over more reliable and available contraception options owing to health system and societal barriers.^5^ Female university students described stigma, fear of being perceived as promiscuous, limited awareness of where to obtain specific products, and the absence of youth-friendly services as main obstacles to accessing HIV and SRH care; highlighting the need for differentiated service delivery methods and youth-friendly spaces.^6^

Hair salons could serve as an innovative and easily accessible community setting to engage young women, including female students, and help overcome certain barriers to SRH services. We found lower acceptability of HIV/SRH services at hair salons than reported in similar studies in neighbouring South Africa^8,9^ and Lesotho.^10^ In South Africa, a research group conducted a pilot study evaluating the uptake of PrEP and contraception through integrated SRH services for women in hair salons.^8^ About 49% of participants accepted salon-based PrEP and 89% accepted salon-based contraception. These services were acceptable to women at high risk or HIV, STIs, and unintended pregnancies. Previously, the same research group had assessed the acceptability and feasibility of providing SRH services in salons through a cross-sectional study, finding high acceptability among both clients and stylists.^9^ Similarly, in a survey conducted in Lesotho, acceptability ranged from 83% to 94%.^10^

Comparisons between our findings and previous evidence should be interpreted with caution, as the studies differed in design and setting. The South African study was a pilot in which services were offered directly, and uptake was measured. Moreover, the service model involved a nurse delivering the services in an adjacent room of the hair salon. In contrast, the study from Lesotho only assessed acceptability (using the same questionnaire as in our study), and suggested delivering the services by trained hair stylists directly. In addition, several contextual factors may help explain differences across settings. HIV prevalence is lower in Zambia than in South Africa and Lesotho, which may reduce the perceived need for such services. Service availability may also influence acceptability. For example, emergency contraception was among the least accepted services in hair salons and was already readily available on campus. In contrast, oral HIV self-testing had higher acceptability, although it was also available on campus, possibly because accessing it required students to visit the campus medical services.

Our exploratory analysis showed that higher HIV acquisition risk increases the acceptability for HIV services in hair salons. This finding suggests that this model of care could be particularly promising for women at higher risk of HIV acquisition. In line with this, the South African study reported that the salon-based model reached young women with risk factors for HIV acquisition.^8^ However, our results are exploratory, given the cross-sectional nature of the data (possible reverse causality) and the potential for unmeasured confounding.

Moreover, due to the remote survey recruitment, the questionnaire did not capture participants’ HIV status or whether they were already using HIV/SRH services.

## Conclusion

Almost half of female students at the university were at increased risk of HIV acquisition, highlighting the need for innovative approaches to improve access to HIV and SRH services in this population. Acceptability of hair salon-based HIV and SRH services was lower than in other similar studies, but may still be promising for selected services, particularly among students at higher HIV acquisition risk. Future work should explore how the model could be adapted to students’ needs, preferences, and the existing service landscape.

## Data Availability

The dataset generated and analysed during the current study is available in the Zenodo repository at https://doi.org/10.5281/zenodo.21527174. The analysis code is available on OSF at https://doi.org/10.17605/OSF.IO/CFDZK.

https://doi.org/10.5281/zenodo.21527174

## List of abbreviations

PrEP: pre-exposure prophylaxis
SRH: sexual and reproductive health
STI: Sexually transmitted infection

## Declarations

### Ethics approval and consent to participate

The study was approved by relevant institutional Review Boards in Zambia (UNZBREC no. 5860-2024; NHRA-1686/07/11/2024) and the USA (COMIRB 22-1151). All participants provided consent prior to participation.

### Consent for publication

Not applicable.

### Authors’ contributions

MC, MK, KH and AA conceptualized the study. KH and OM obtained the funding for the overarching randomized trial, on which this survey is based on. AA, LN, TH, and OM drafted the survey questionnaire over several iterations, reviewed by KH. LN, TH, OM led the survey implementation, supervised by KH. MC conducted the analysis, supervised by AA. MC drafted the first version of the manuscript. All named authors reviewed the draft manuscript, provided input, and approved the final version.

## Supplementary material

**Supplementary 1.**
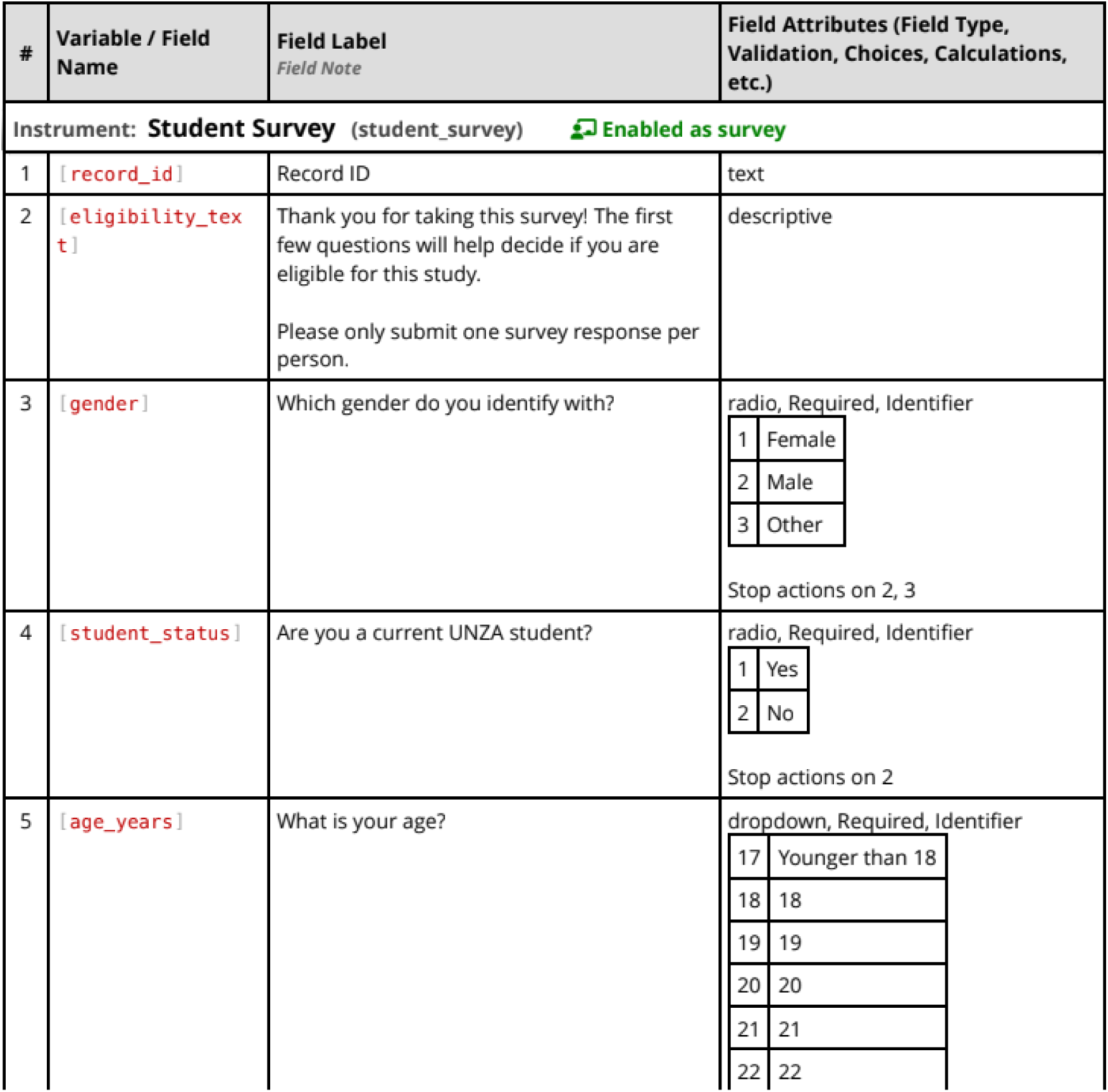
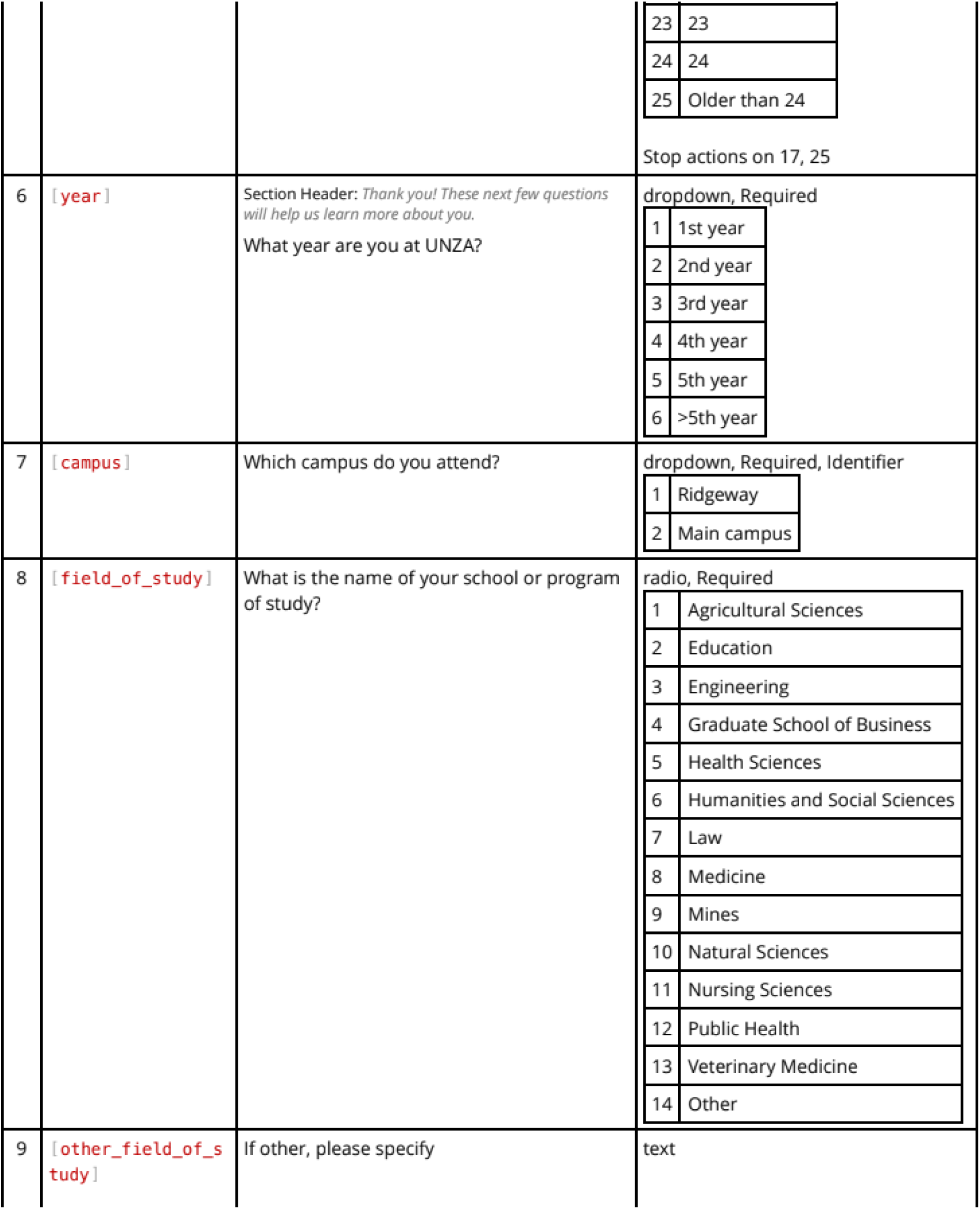
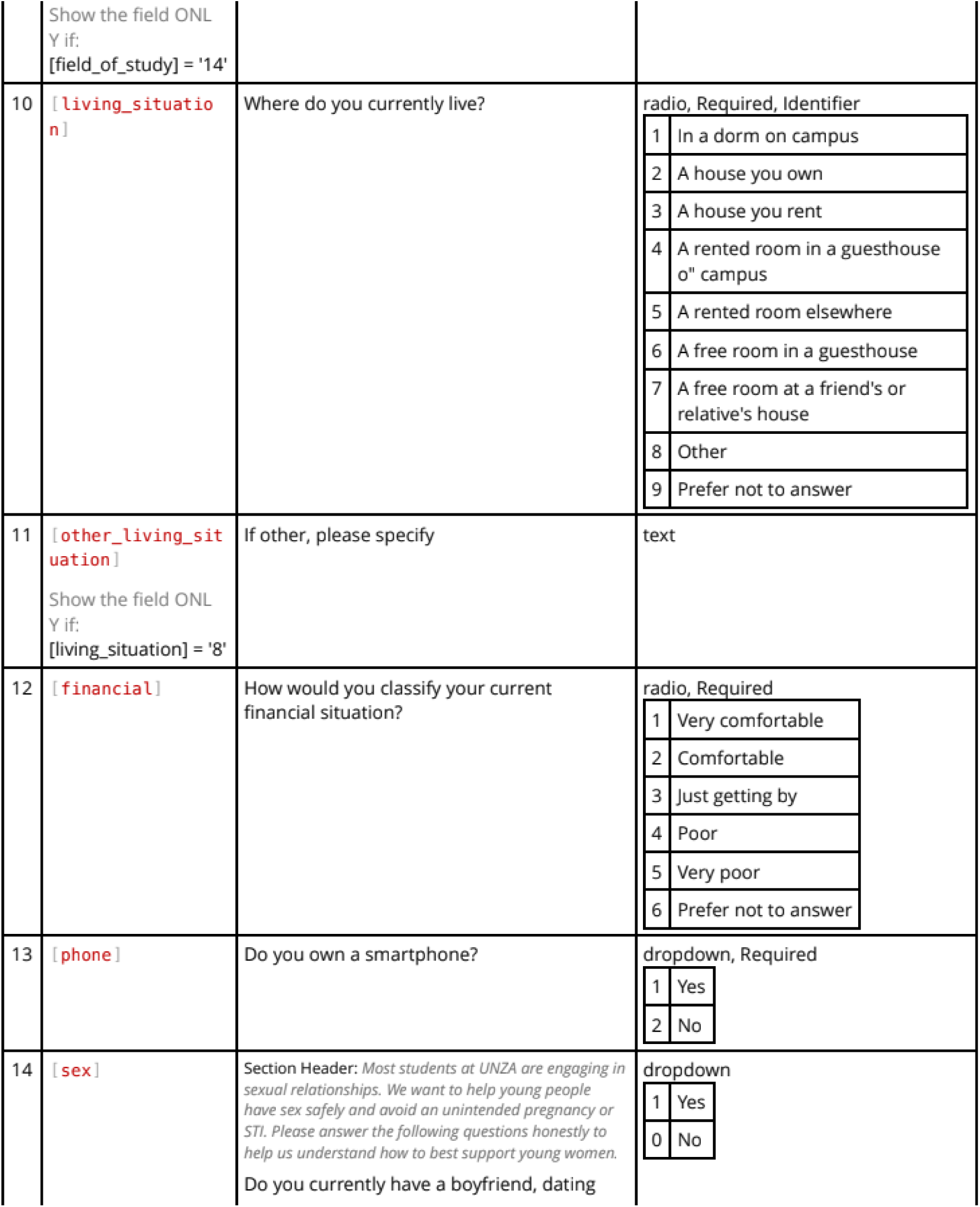
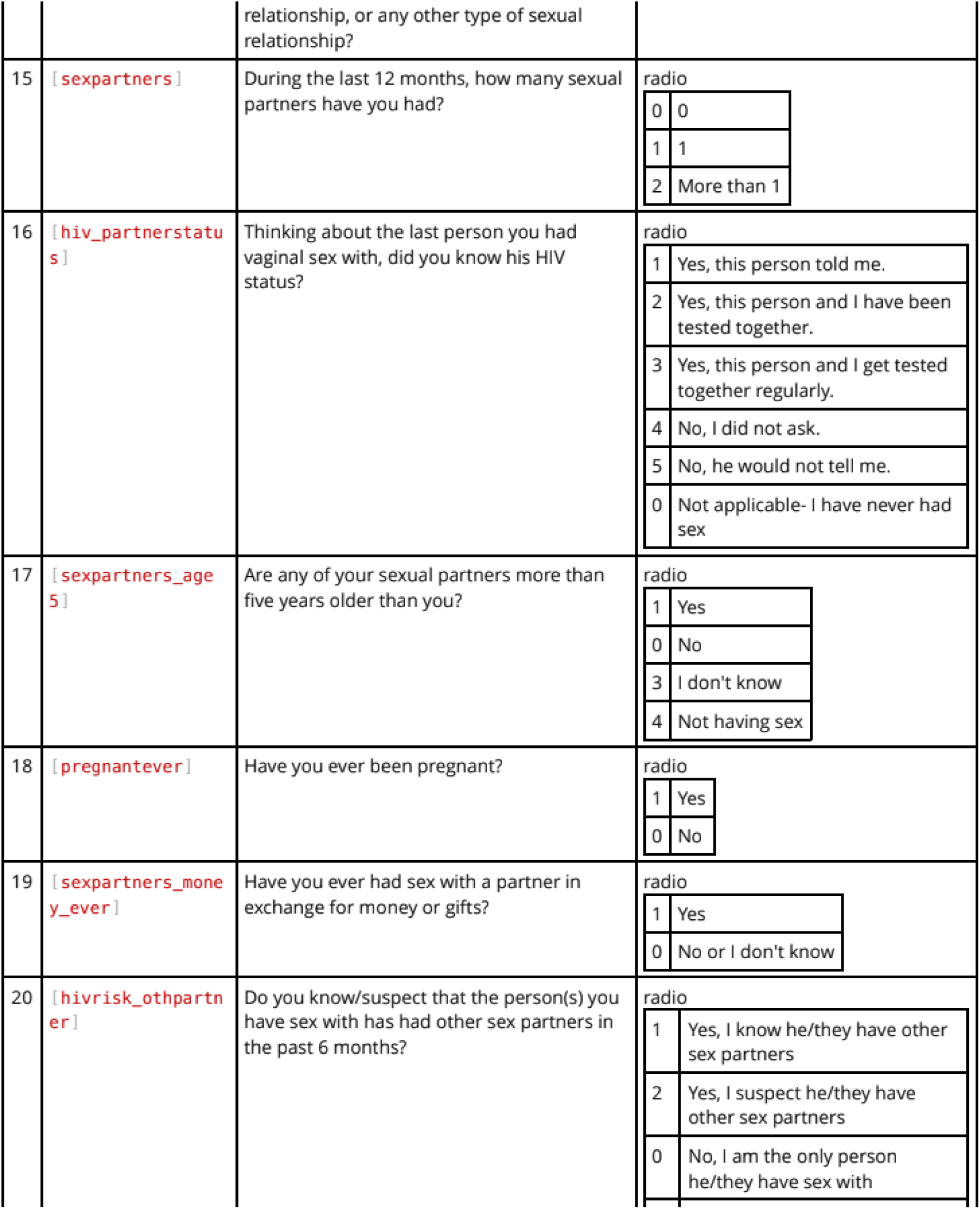
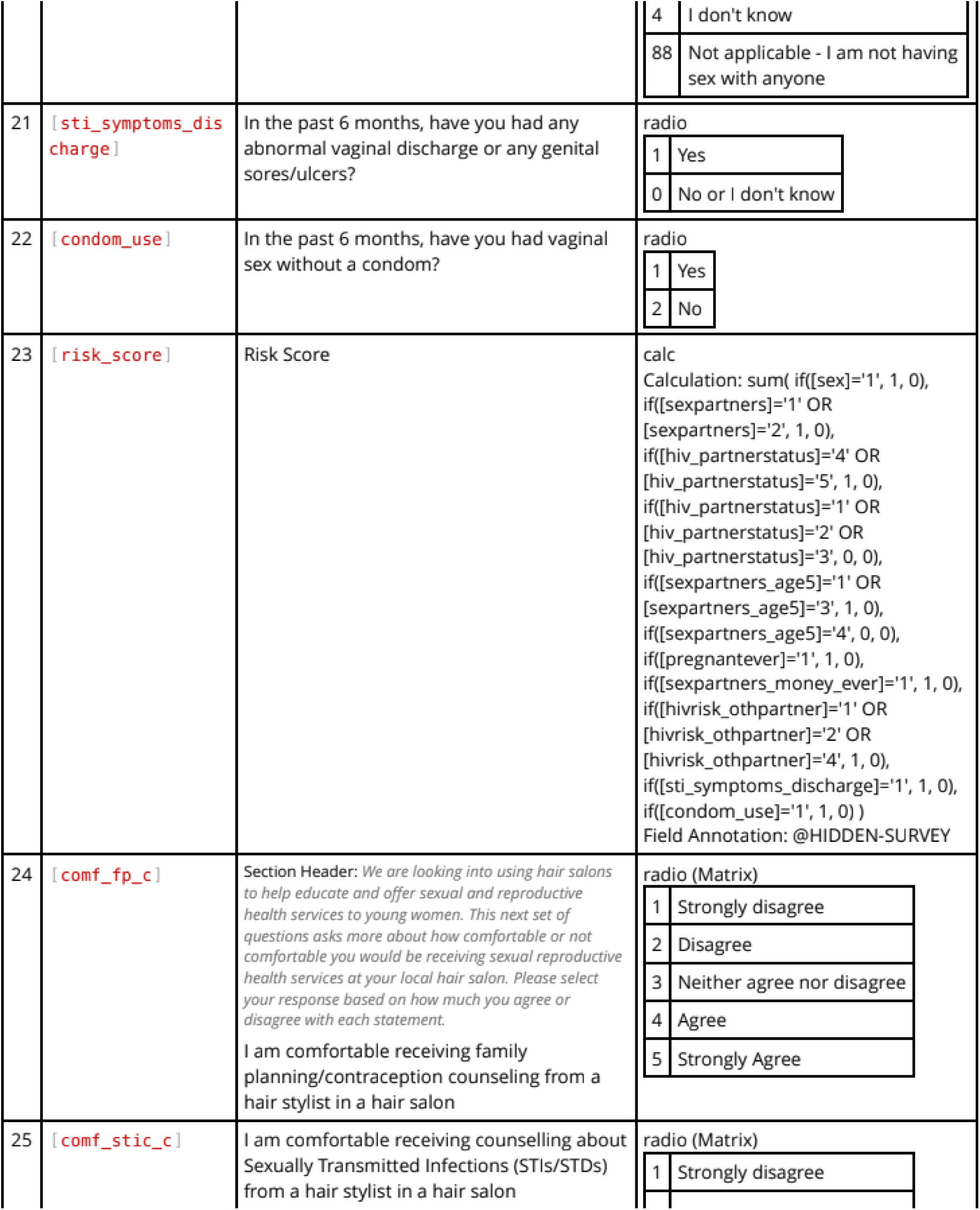
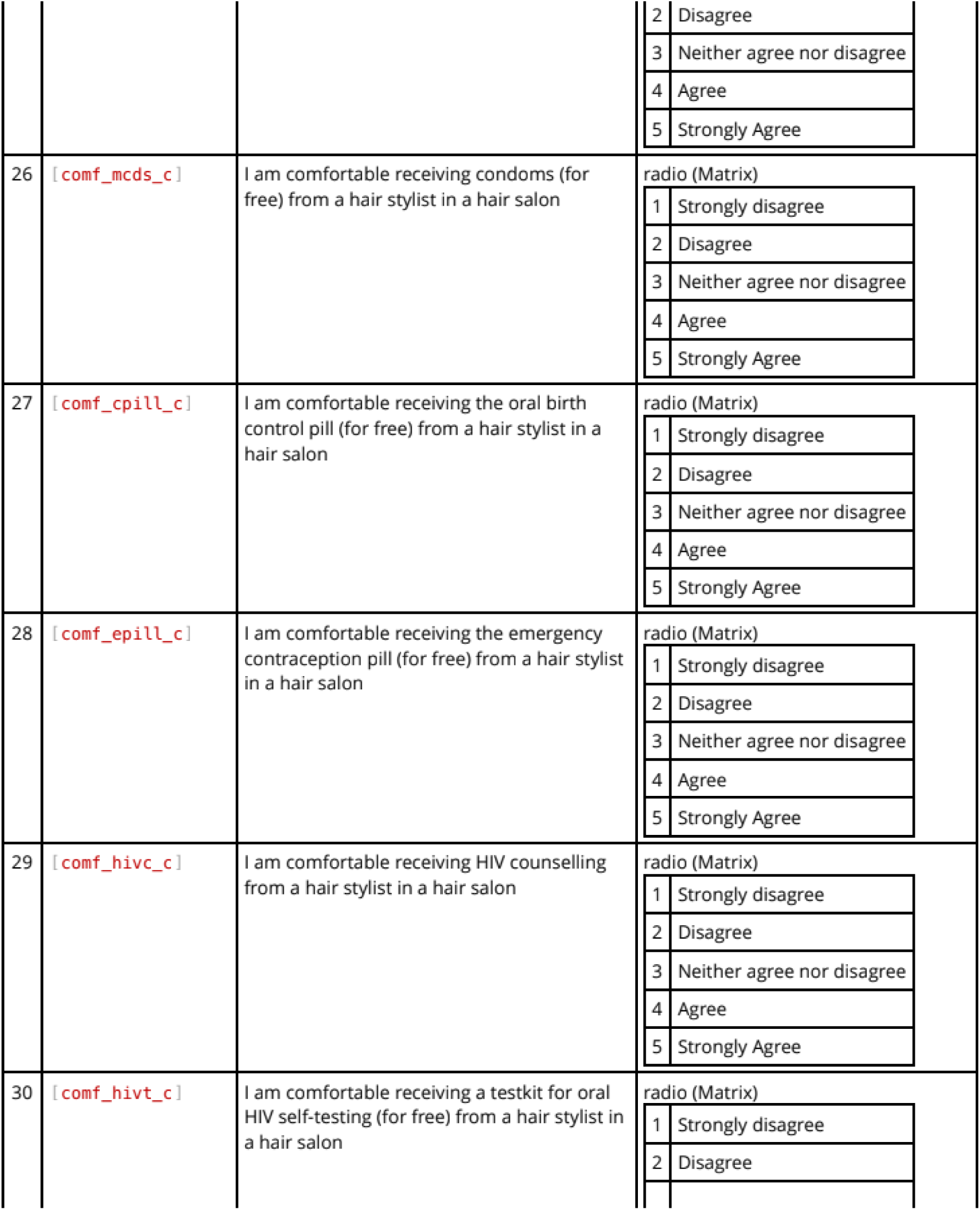
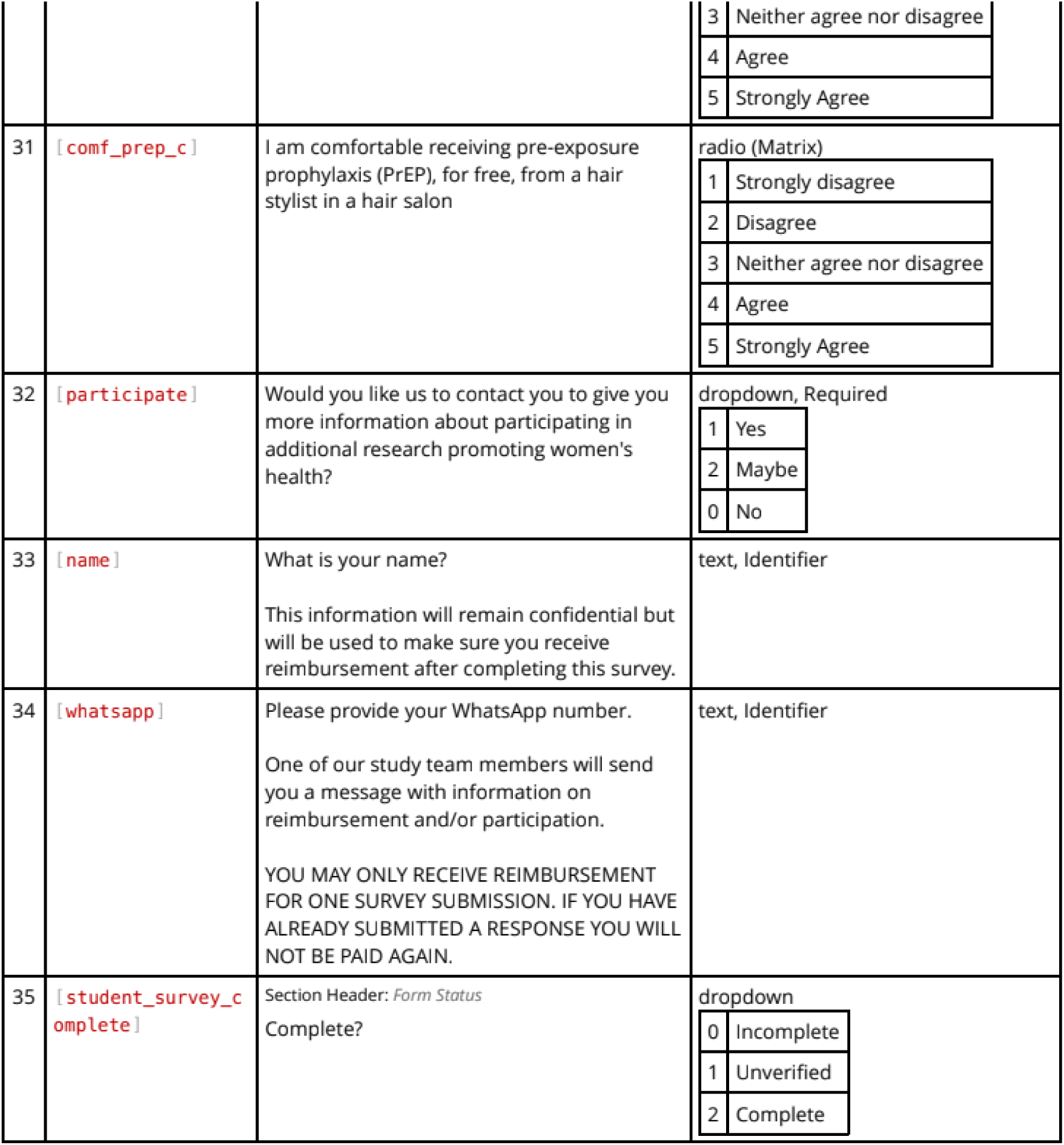
Questionnaire.

**Supplementary 2.**
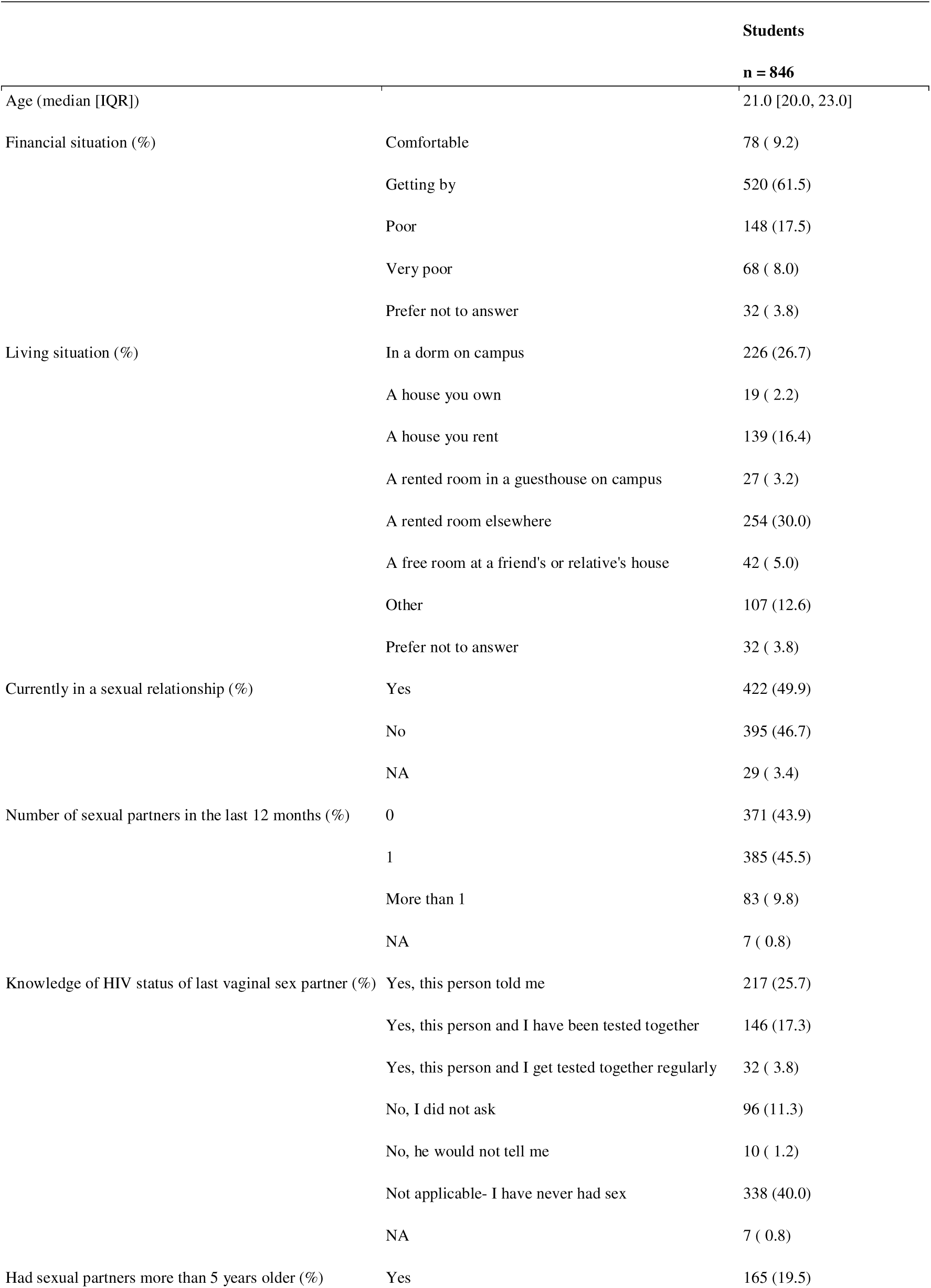
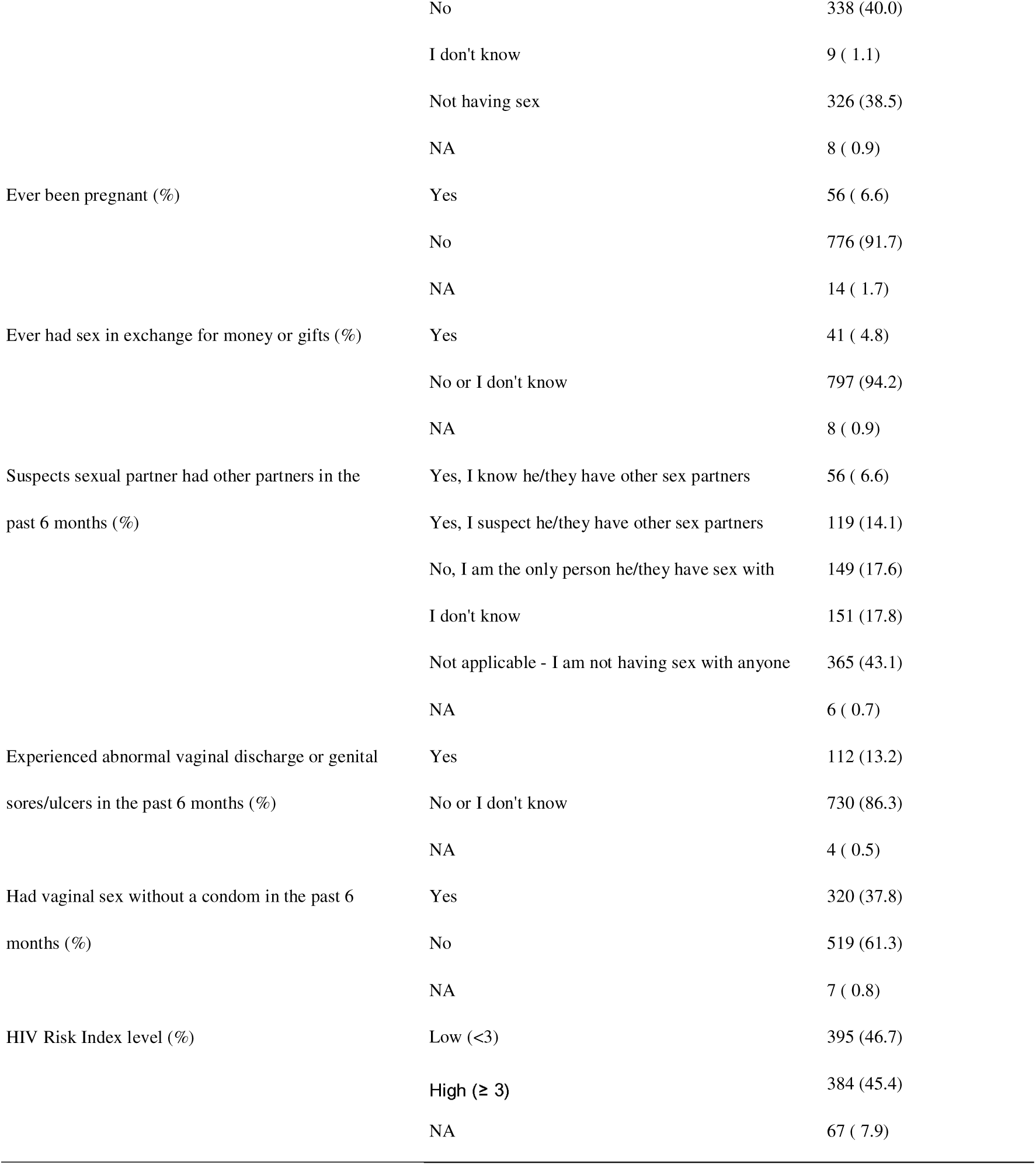
Students’ baseline characteristics and risk factors for HIV acquisition.

**Supplementary 3.** Acceptability of students receiving HIV and SRH services in hair salons.

| n = 846 | Strongly disagree | Disagree | Neutral | Agree | Strongly agree | Missing |
| --- | --- | --- | --- | --- | --- | --- |
| HIV counselling (%) | 152 (18) | 187 (22) | 120 (14) | 272 (32) | 101 (12) | 14 (2) |
| Oral HIV self-testing (%) | 160 (19) | 193 (23) | 110 (13) | 262 (31) | 108 (13) | 13 (2) |
| HIV pre-exposure prophylaxis (%) | 189 (22) | 258 (30) | 137 (16) | 170 (20) | 79 (9) | 13 (2) |
| Family planning counselling (%) | 241 (28) | 155 (18) | 126 (15) | 216 (26) | 99 (12) | 9 (1) |
| Sexually transmitted infection counselling (%) | 123 (15) | 180 (21) | 114 (13) | 299 (35) | 120 (14) | 10 (1) |
| Condoms (%) | 159 (19) | 238 (28) | 129 (15) | 220 (26) | 88 (10) | 12 (1) |
| Birth control pill (%) | 199 (24) | 285 (34) | 126 (15) | 162 (19) | 62 (7) | 12 (1) |
| Emergency contraceptive pill (%) | 194 (23) | 256 (30) | 130 (15) | 181 (21) | 71 (8) | 14 (2) |

